# Functional Neural Networks Stratify the Parkinson’s Disease Patients Across the Spectrum of Cognitive Impairment

**DOI:** 10.1101/2023.05.13.23289935

**Authors:** Farzin Hajebrahimi, Miray Budak, Mevhibe Sarıcaoğlu, Lütfü Hanoğlu, Zübeyir Bayraktaroğlu, Süleyman Yıldırım

## Abstract

Cognitive Impairment (CI) in Parkinson’s disease (PD) is one of the important non-motor symptoms that can begin even before the motor symptoms manifest. As the disease progresses into advance stages, however, virtually all patients suffer from cognitive decline. PD Patients hypothetically progress across PD with no CI (PD-NC), Mild Cognitive Impairment (PD-MCI), and PD dementia (PDD). The CI symptoms in PD are linked to different brain regions including dysfunctional subcortical regions and poorly elucidated neural pathways. However, it is still unknown how functional dysregulation in some brain regions correlates to CI progression in PD. Recently, rsfMRI has been shown to be a promising neuroimaging technique that can enable discovery of CI biomarkers in PD. Here, we investigated the differences in the clinical measures and the resting-state Functional Connectivity (FC) of three CI subtypes of PD. We included a total of 114 participants, (26 PD-NC, 32 PD-MCI, 31 PDD, and 26 Healthy Controls (HC), and performed intra- and inter-network FC analysis together with comprehensive clinical cognitive assessment. Our results showed the importance of several neural networks including Default Mode Network (DMN), Frontoparietal Network (FPN), Dorsal Attention Network (DAN), and Visual Network (VN) and their inter-intra network FC distinguishing between PD-MCI and PDD. Additionally, our results showed the importance of Sensory Motor Network (SMN), VN, DMN, and Salience Network (SN) in the discriminating PD-NC from PDD. Finally, in comparison to HC, we found DMN, FPN, VN, and SN as important networks for further differential diagnosis of CI subtypes of PD. We propose that resting state networks can be used in stratifying the CI subtypes of PD patients in the clinic.

## Introduction

Parkinson’s disease (PD) is a common chronic and progressive neurodegenerative disease characterized by the loss of dopaminergic neurons in the substantia nigra and α-synucleinopathy (Aarsland & Kramberger, 2015; Göttlich et al., 2013). The dysfunctional cortico-striatal-thalamic-cortical loop in PD, causes cardinal motor symptoms such as tremor, akinesia, and rigidity (Jankovic, 2008). Moreover, multiple neurotransmitter deficiencies result in heterogeneity in the clinical phenotype, associating with a wide spectrum of motor and non-motor clinical symptoms (Aarsland & Kramberger, 2015; Lee et al., 2022). The symptoms and progression of PD vary greatly from person to person (Fereshtehnejad et al., 2017), in addition to the important role of clinical phenotypes, age of onset, disease severity, or neuropathological changes in PD progression (Krüger et al., 2017). On the other hand, alterations in many functional brain networks are one of the potential underlying mechanisms in the relationship between neuropathology and behavioral outcomes in PD (Gratton et al., 2018). The widespread symptoms and functional neuropathology in PD emphasize the need to study PD functional networks and their alterations in relation to each other (Filippi et al., 2018).

Non-motor symptoms have recently been shown to play an increasingly important role in the clinical heterogeneity of PD (Mu et al., 2017). One of the major non-motor symptoms is cognitive impairment (CI), which includes working memory deficits, planning, visuospatial, and set-shifting problems (Khoo et al., 2013). CI can start years before the motor manifestations in PD patients (Aarsland et al., 2017). Importantly, the presence of mild cognitive impairment (MCI) in PD (PD-MCI) is a transitional state between PD with normal cognition (PD-NC) and PD with dementia (PDD) and has been introduced as an independent predictor for PDD (Aarsland & Kurz, 2010). Therefore, early detection of the CI and consequently the PD-MCI would play an integral role in the course of PD diagnosis and treatment. Of note, the symptoms of CI in PD are linked to changes in a number of different brain regions via neural pathways, in addition to being the result of dysfunctional subcortical regions (Suo et al., 2022). However, it is still unknown how functional dysregulation correlates to disease progression (Filippi et al., 2021).

Recently, advanced neuroimaging techniques have been instrumented in identifying the alterations in functional connections associated with PD and providing a deeper understanding of the disease (Caspers et al., 2021). There is emerging evidence for using the functional Magnetic Resonance Imaging (fMRI) findings as biomarkers of CI (i.e., PD-NC, PD-MCI and PDD) in PD (Hou & Shang, 2022; Rektorova et al., 2014). Resting-state functional MRI (rs-fMRI) is a relatively new technique for assessing spontaneous or intrinsic neural activity, which measures slow oscillations in the blood-oxygen-level dependent (BOLD) signal while subjects are awake and not performing any specific task (Tahmasian et al., 2017). The rs-fMRI enables identification of brain networks associated with symptoms of the disease (Filippi et al., 2021) and can be a critical tool for investigating the causal factors of CI in PD and in the process of diagnosis (K. Li et al., 2018). The altered functional connectivity (FC) of the default mode (DMN), dorsal-attention (DAN), fronto-parietal (FPN), salience (SN), and related visual networks (VN) are among the previously reported rs-fMRI hallmarks in PD patients with CI (Filippi et al., 2018). Furthermore, the rs-fMRI findings in studies of CI subtypes of PD (i.e., patients with PD-NC, PD-MCI and PDD) based on the predefined cognitive levels have shown differences in the FC, but there is not a clear consensus over differentiating neural correlates across the spectrum of PD-CI (Wolters et al., 2019). In this context, there are only a few studies that have included all three clinically pre-defined CI subtypes of PD (i.e., patients with PD-NC, PD-MCI and PDD) in their analysis, and specifically, the number of PDD patients included in these studies is relatively low. Additionally, due to the heterogeneity of the analysis methods used to investigate brain FC, the need to implement methods that can be further used in clinical diagnosis and individualized medicine and also distinguish different subtypes of CI in the progression of PD is highly warranted in recent years. Therefore, there is a need for studies that include all three CI subtypes of PD cognitive phenotypes, specifically patients with PDD, to better understand the FC and clinical differences between the CI subtypes of PD. To address these problems, we investigated the differences in the clinical outcomes and the resting-state FC of all three CI subtypes of PD and compared them among each other and also with HC. We hypothesized that FC shifts in association with CI progression in PD and includes the discriminatory factors for diagnosing and predicting subtypes of CI in PD. The purpose of this study was to investigate clinical and inter- and intra-network differences in the CI subtypes of PD by reporting the differences between three CI subtypes and better defining the characteristics of the PD-MCI and PDD subtypes, showing the transition between CI subtypes, and providing a link between neural correlates and clinical outcomes. This paper shows how the three CI subtypes of PD differ from each other in terms of clinical outcomes and resting-state FC, as well as how they differ from HC. We show that the results of rs-fMRI networks, in conjunction with clinical factors related to cognitive outcomes, can be useful in distinguishing between CI subtypes of PD. This can hopefully highlight the treatment plans based on the differences between the CI subtypes of PD.

## Material and Methods

### Study Design and Participants

The ethics committee of the Istanbul Medipol University approved this study with authorization number 10840098-604.01.01-E.3958. We used a cross sectional study design. We first assessed 138 participants for their eligibility in the study. PD patients were recruited in the neurology clinic at the Istanbul Medipol University Medipol Mega Hospital (Bagcilar, Istanbul). We screened these participants based on the inclusion and exclusion criteria (see below) and included 94 PD patients and 26 HC participants in the initial evaluation. All participants gave their informed consent in accordance with the principles of the Declaration of Helsinki. We completed the data collection between 2018 and 2022. An experienced neurologist (co-author LH) evaluated the patients and gave the clinical diagnosis of PD within the framework of Brain Bank criteria (Gelb et al., 1999), followed by the recommendations of the Movement Disorder Society (MDS) regarding CI in PD (Dubois et al., 2007; Emre et al., 2007; Pourzinal et al., 2022).

### Inclusion Criteria

Inclusion criteria for the PD patients were the diagnosis of PD, receiving stable anti-parkinsonian medication for at least 1 month, and aged between 45-90 years old.

### Exclusion Criteria

Exclusion Criteria were the history of unstable medical treatment, pyramidal findings, cerebellar involvement, gaze palsy, and autonomic dysfunction, receiving device-assisted therapy, and contraindications for MRI scanning.

### Clinical Evaluation

After a neurological examination, we performed the comprehensive neuropsychological testing and fMRI scans for all the participants. Besides screening tests for PD patients, including the Hoehn & Yahr test, the Unified Parkinson’s Disease Rating Scale-Motor (UPDRS-III) and demographic screening, we performed a precise cognitive assessment by means of a Neuropsychological Test Battery. A multidisciplinary team of neurologists, neuropsychologists, and physical therapists conducted the overall assessment of the patients.

### Cognitive Assessment

We used the Mini-Mental State Examination (MMSE) to assess global cognition (Güngen et al., 2002). We used the Digit Span Test to evaluate attention functions (Kurt et al., 2011). We then used the Wechsler Memory Scale (WMS) to evaluate memory functions including the WMS Visual Reproduction and Recognition Test (Immediate and Delayed) (KARAKAŞ et al., 1999), and Oktem Verbal Memory Process Test (Turkish Verbal Memory Test: SBST) (Bosgelmez et al., 2015). We evaluated the language skills by the Boston Naming Test (Soylu & Cangöz, 2018). We evaluated visual and perceptual functions by Judgement of the Line Orientation Test (Spencer et al., 2013) and Benton Facial Recognition Test (Schretlen et al., 2001). We evaluated executive functions by Stroop Test (KARAKAŞ et al., 1999), and the Clock Drawing Test (Shulman, 2000). Finally, we evaluated apathy with the Apathy Evaluation Scale (AES) (Marin et al., 1991). We used the Turkish versions of the memory, language, and executive functions tests.

### Functional Magnetic Resonance Imaging Protocol

### Data Acquisition

We conducted structural and functional MRI scans at the Istanbul Medipol University Medipol Mega Research and Training Hospital in the Radiology clinic (Bagcilar, Istanbul). To conduct rs-fMRI we used a 32-channel head coil using standard sequences. We explained the nature of the MR environment to all participants, and trained them to act accordingly, with regard to the safety of patients with movement disorders (Van Dijk et al., 2012). Similar to our previous studies in patients with neurodegenerative diseases (Budak et al., 2022; Hajebrahimi et al., 2022), to get the benefit of the highest level of participants’ alertness, was planned the functional scan as the earliest in the imaging queue, just after the localizer protocol: (1) localizer, (2) resting-state fMRI, (3) fieldmap, (4) T1 weighted structural and (5) T2 weighted structural scans. We scanned the patients during their “ON” period. We asked the participants to fix their gaze on a spot inside the scanner, during resting-state data acquisitions, while keeping their eyes open and not thinking about anything particular or rhythmic (praying, counting, tapping, etc.), and instructed to stay still as much as possible. We suggested the participants close their eyes and rest during the structural scans. To minimize movement artifacts, we stabilized the patients’ heads with spongy pads inserted between the sides of the head and the MR coil. The imaging process took approximately 30 minutes to complete, including preparation time.

We acquired the structural T1 and T2 images in the sagittal plane (TR/TE: 8.1/3.7), FOV 256×256×190 mm (FH×AP×RL), with the voxel size of 1 × 1 × 1 mm3. The functional scan parameters were 300 volumes (TR/TE: 2230/30 ms, FA 77°), FOV 240×240×140 mm (RL×AP×FH), voxel size of 3 × 3 × 4 mm3, and 35 slices. These parameters, including scanning protocols and their order, were the same in all participants except for a difference in TR and volume of functional scans due to a scanner upgrade while the study was in progress (TR:2000 ms, # of volumes: 341). To eliminate the effects of this difference in the group comparisons and network analysis, we included covariates of no interest in the GLM design during the analysis steps. Additionally, we included age, brain volume (in mm^3^-normalized to MNI), and L-dopa equivalent daily dose (LEDD) as covariates of no interest in the GLM design (Tahmasian et al., 2017).

### Analysis

### fMRI Data Analysis

We used FMRIB FSL software package for data preprocessing and subsequent functional network analysis (Jenkinson et al., 2002). We first convert DICOM files into NIFTI format using the dcm2niix tool. (X. Li et al., 2016). Next, we used the fsl_anat script for brain extraction from the NIFTI images. After brain extraction, we used the tools supplied in the FSL FEAT interface for motion correction and smoothing with a Gaussian kernel of 5 mm full width at half maximum (FWHM) in each voxel. To filter the functional data, we used a 150s high-pass filter. We used the FSL MCFLIRT algorithm to correct the head motions between dynamic scans (Jenkinson et al. 2002). We spatially normalized the functional and anatomical data to MNI152 for group comparisons; in three steps using the FSL FLIRT and FNIRT linear and non-linear registration tools, respectively. Using limited transformations, we first matched the functional brain images to the participants’ own high-resolution anatomical images (6 DOF). Next, we matched the participants’ high-resolution anatomical images with MNI152 standard brain images, utilizing 12 DOFs and non-linear transformations in the second step. The distortion resolution was ten millimeters. In the third step, using matrices from previous transformations, we mapped low-resolution functional images to the MNI152 brain. In a separate step, we processed each participant’s functional data in the native space into an exploratory Independent Component Analysis (ICA) using the FSL MELODIC tool to determine movement, physiological (heart, respiration, etc.), and other imaging artifacts. By inspecting signal features of ICA components such as spatial distribution, frequency spectrum, and temporal fluctuation, we manually hand-classified the components and filtered those compatible with the noise from the functional data using the fsl_regfilt command (Griffanti et al., 2017). We then transformed the functional data into the standard space after artifact removal in the native space.

### Independent Component Analysis and Dual Regression

For group comparison, we also utilized the FSL MELODIC tool to perform a group-level ICA. For this reason, we used a temporal concatenation approach in FSL MELODIC to perform group-ICA with 20 components on the preprocessed artifact-free data (C. F. Beckmann & Smith, 2004). Afterwards, we evaluated each component’s spatial correlation with resting-state networks and labeled the components consistent with the resting-state networks explained elsewhere (Smith et al., 2009). Next, we used FSL Dual Regression and to first regress the group-level ICs into all subjects’ data and extract timeseries of each of the 20 components and in the second regression, extracting timeseries into the same resting state data of the participants to calculate the subject-specific spatial maps of each ICs (C. Beckmann et al., 2009; Nickerson et al., 2017). We finally calculated z-stat images for each of the subject-specific spatial maps. For the group comparisons, we designed a GLM for a one-way ANOVA (4 groups × 1 time). We included TR, volume, age, brain volume (in mm3-normalized to MNI) and L-dopa equivalent daily dose (LEDD) as covariates of no interest in our group-level GLM design. We used the FSL randomize tool for non-parametric inference with 5000 random permutations to examine the significance of the differences. We corrected multiple comparisons using cluster-based Threshold-Free Cluster Enhancement (TFCE) and considered the results significant at p<0.05.

### Network Connectivity Analysis

We used the tools implemented in FSLNets (http://fsl.fmrib.ox.ac.uk/fsl/fslwiki/FSLNets) for inter-network connectivity analysis. For this reason and to better compare the results of network analysis with the literature, we performed the network analysis utilizing a functional atlas. We used the Schaefer 2018 atlas with 100 parcellations, which projects on the Yeo 17 Networks over FSLMNI152 (Schaefer et al., 2018). Afterwards, we extracted subject-specific time series using the first stage of Dual Regression to shape the inter-network connectivity matrices. We calculated full and partial correlations between 100 nodes. While full correlation can demonstrate indirect connections between two specific nodes, in partial correlation, other nodes are controlled to calculate the correlation between two specific nodes, which therefore may provide better information regarding the direct connectivity (Bijsterbosch et al., 2017). After obtaining the full and partial correlation matrices across all subjects, we extracted the full and partial correlation matrices of each group separately. Consequently, we prepared the hierarchical clustering for each of the described groups by utilizing the nets_hierarchy tool in MATLAB (a total of 4 hierarchical clustering dendrograms as HC, PD-NC, PD-MCI, and PDD). Next, we used the partial and full correlation matrices in all 4 groups to create a connectome to compare the results with hierarchical clusterings. Finally, we used the same GLM design explained above to compare the groups regarding inter-network connectivity. Here, we fed 100 nodes into the GLM design and included previously described covariates (i.e. TR, number of volumes, age, brain volume and LEDD) in the GLM design as covariates of no interest. The cross-subject GLM on the partial correlation netmats (FSL-randomize with 5000 permutations) gave uncorrected and corrected p values as output (i.e., corrected for multiple comparisons with TFCE across the 100x100 netmat elements). We extracted the corrected p values to report the differences in the network organization between groups. Furthermore, we reported contrasts and their related significant nodes. To better visualize the differences between groups related to the inter-network connectivity, we compared the connectograms of groups and showed multiple significant nodes with their connections (here, partial connections) to the entire network. Of note, we used those nodes that were significant more than once (at least 2) in the group comparison to visualize the difference. Finally, to better conceptualize how our results explain the CI progression across the cognitive spectrum of the PD, we prepared an overall continuum map to better show the transition between different CI subtypes of the disease, merging all the results coming out of the group-level ICA and network FC comparison.

### Statistical Analysis

We used IBM SPSS (Statistical Package for Social Science) version 25.0 for statistical analysis. We presented the mean, standard deviation, and percentage values in the descriptive statistics of the data. We measured the normal distribution of the variables with the Kolmogorov Smirnov test. Next, we evaluated the nominal data of the independent variables with the Chi-Square test, and the numerical data with the One-Way ANOVA test. The significance value was accepted as p<0.05.

### Sample Size

The sample size was determined using the "G*power sample size calculator" (Faul et al., 2007). The sample size was calculated as 112 subjects using “ANCOVA: Fixed effects, main effects, and interaction” design for four groups with a power of 95% (α=0.05, β=0.95, λ=17.92, F=2.68) and an effect size of 0.40.

## Results

From the included participants, 6 PD patients refused to complete the study and were excluded. We included 88 patients with PD in the study and allocated them into three groups as the PD-Normal Cognition (PD-NC) Group (n = 25), the PD-Mild Cognitive Impairment (PD-MCI) Group (n = 32), and the PD-Dementia (PDD) Group (n = 31) according to the Clinical Dementia Rating score (CDR). We also included 26 Healthy Controls (HC) in the study (Fig. 1).

**Figure 1.**
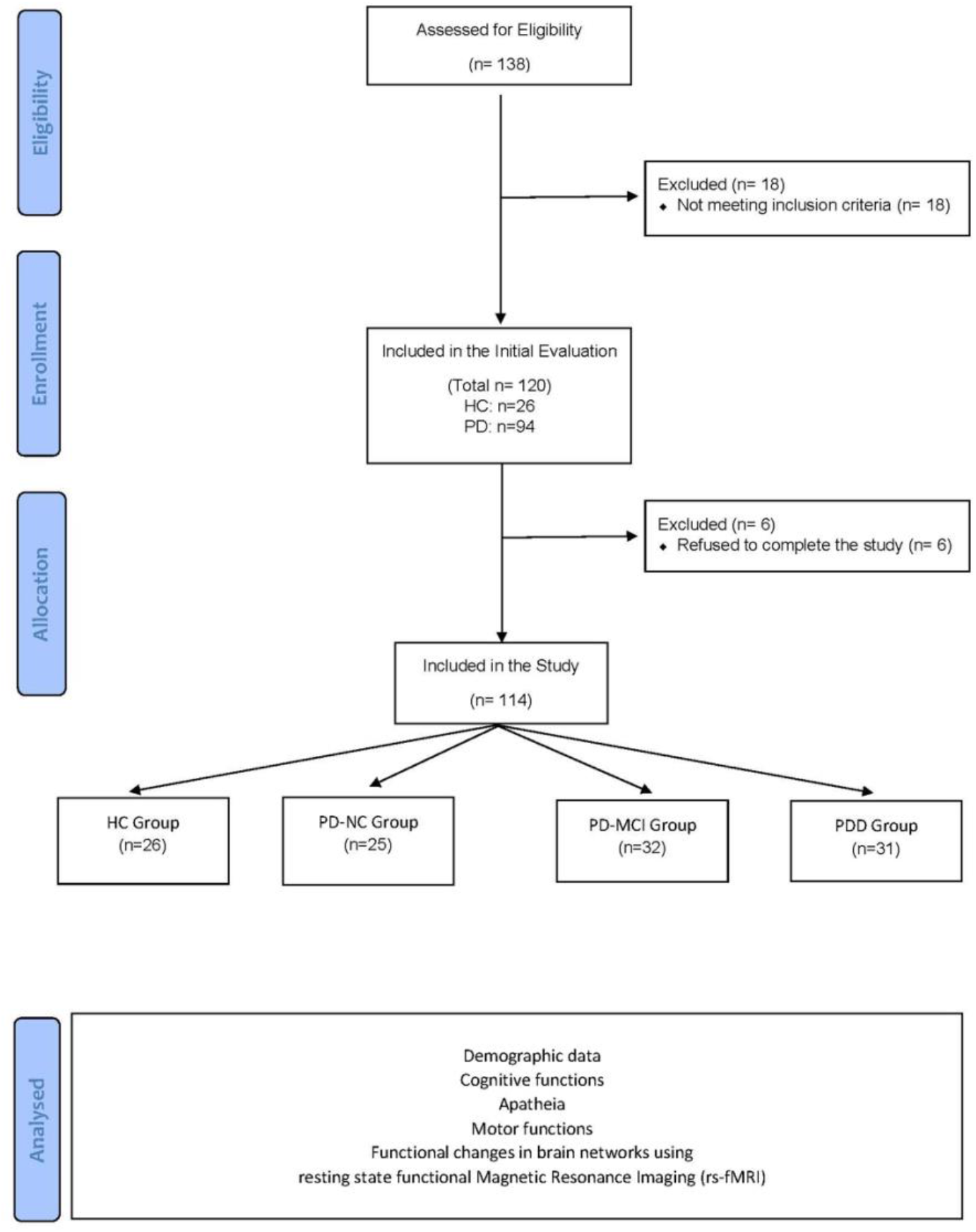
- Flowchart of the study

### Cognitive Assessment Results

The demographic and clinical variables and their statistical comparisons are shown in Table 1. 49% (56/114) of the participants were female. Patients with PDD were significantly older (71.77 ± 8.87), had a longer duration of the disease (101.88 ± 45.20, months), had worse LEDD, UM-PDHQ and UPDRS-III compared with the PD-NC, PD-MCI, and HC groups (p<0.05). Between-group cognitive status is shown in Table 2. There were statistically significant differences in all parameters between groups (p<0.05). As expected, the PDD group performed worse in cognitive assessments when compared to PD-NC, PD-MCI and HC groups. There was statistically significant deterioration in terms of all clinical parameters between all groups (p<0.001). Fig. 2 shows the progression of clinical cognitive outcomes from HC to PD-NC, PD-MCI and PDD. In all clinical outcomes, there was a significant gradual decrease in scores from HC to PD-NC, from PD-NC to PD-MCI, and from PD-MCI to PDD (p<0.001).

**Table 1.** Distribution of demographic data

|  |  | HC<br>(n= 26) | PD-NC<br>(n= 25) | PD-MCI<br>(n= 32) | PDD<br>(n= 31) | Between Group<br>Differences |  |
| --- | --- | --- | --- | --- | --- | --- | --- |
| Age (Avg ± SD) | | 58.19 ± 8.09 | 67.84 ± 9.04 | 67.87 ± 9.94 | 71.77 ± 8.87 | $F_3=11.191$,<br>$p=0.000^*$ | |
| Gender | Female (n / %) | 17 / 65.4 | 6 / 24 | 16 / 50 | 17 / 54.8 | $X_3=9.480$, $p=0.024^*$ | |
|  | Male (n / %) | 9 / 34.6 | 19 / 76 | 16 / 50 | 14 / 45.2 |  |  |
| Education level (Avg ± SD) (years) | | 9.92 ± 5.18 | 6.48 ± 3.87 | 4.12 ± 4.23 | 10.22 ± 11.41 | $F_3=5.146$, $p=0.002^*$ | |
| Dominant<br>hand | Right (n / %) | 26 / 100 | 25 / 100 | 32 / 100 | 31 / 100 | -- | -- |
|  | Left (n / %) | 0 / 0 | 0 / 0 | 0 / 0 | 0 / 0 |  |  |
| Duration of disease (Avg ± SD) (months) | | 0 / 0 | 47.85 ± 32.12 | 69.22 ± 48.39 | 101.88 ± 45.20 | $F_3=33.758$,<br>$p=0.000^*$ | |
| LEDD | | -- | 649.00 ± 393.81 | 750.16 ± 253.98 | 832.12 ± 425.15 | $F_2=1.770$, $p=0.177$ | |
| CDR<br>(n / %) | CDR-0: no cognitive impairment | 26 / 100 | 25 / 100 | 0 / 0 | 0 / 0 | $X_3=222.314$,<br>$p=0.000^*$ | |
|  | CDR-0.5: questionable or very mild dementia | 0 / 0 | 0 / 0 | 0 / 0 | 0 / 0 |  |  |
|  | CDR-1: mild | 0 / 0 | 0 / 0 | 32 / 100 | 0 / 0 |  |  |
|  | CDR-2: moderate | 0 / 0 | 0 / 0 | 0 / 0 | 32 / 100 |  |  |
|  | CDR-3: severe | 0 / 0 | 0 / 0 | 0 / 0 | 0 / 0 |  |  |
| UM-PDHQ (Avg ± SD) | | 0 / 0 | 1.84 ± 0.55 | 1.28 ± 2.42 | 4.48 ± 4.66 | $F_3=10.812$,<br>$p=0.000^*$ | |
| UPDRS (Avg ± SD) | | 0 / 0 | 24.04 ± 18.59 | 29.70 ± 13.94 | 49.35 ± 20.32 | $F_3=47.051$,<br>$p=0.000^*$ | |
| Hoehn-Yahr Scale (Avg ± SD) | | 0.00 ± 0.00 | 2.09 ± 0.76 | 1.79 ± 0.82 | 2.69 ± 0.95 | $F_3=64.343$,<br>$p=0.000^*$ | |

PD: Parkinson's Disease; HC: Healthy Control; PD-NC: PD-Normal Cognition; PD-MCI: PD-Mild Cognitive Impairment; PDD: PD Dementia, Avg: Average, SD: Standard deviation, CDR: Clinical Dementia Rating, L-dopa equivalent daily dose (LEDD), UM-PDHQ: University of Miami Parkinson's disease Hallucinations Questionnaire, UPDRS: Unified Parkinson's Disease Rating Scale. \* $p<0.05$

**Table 2.**
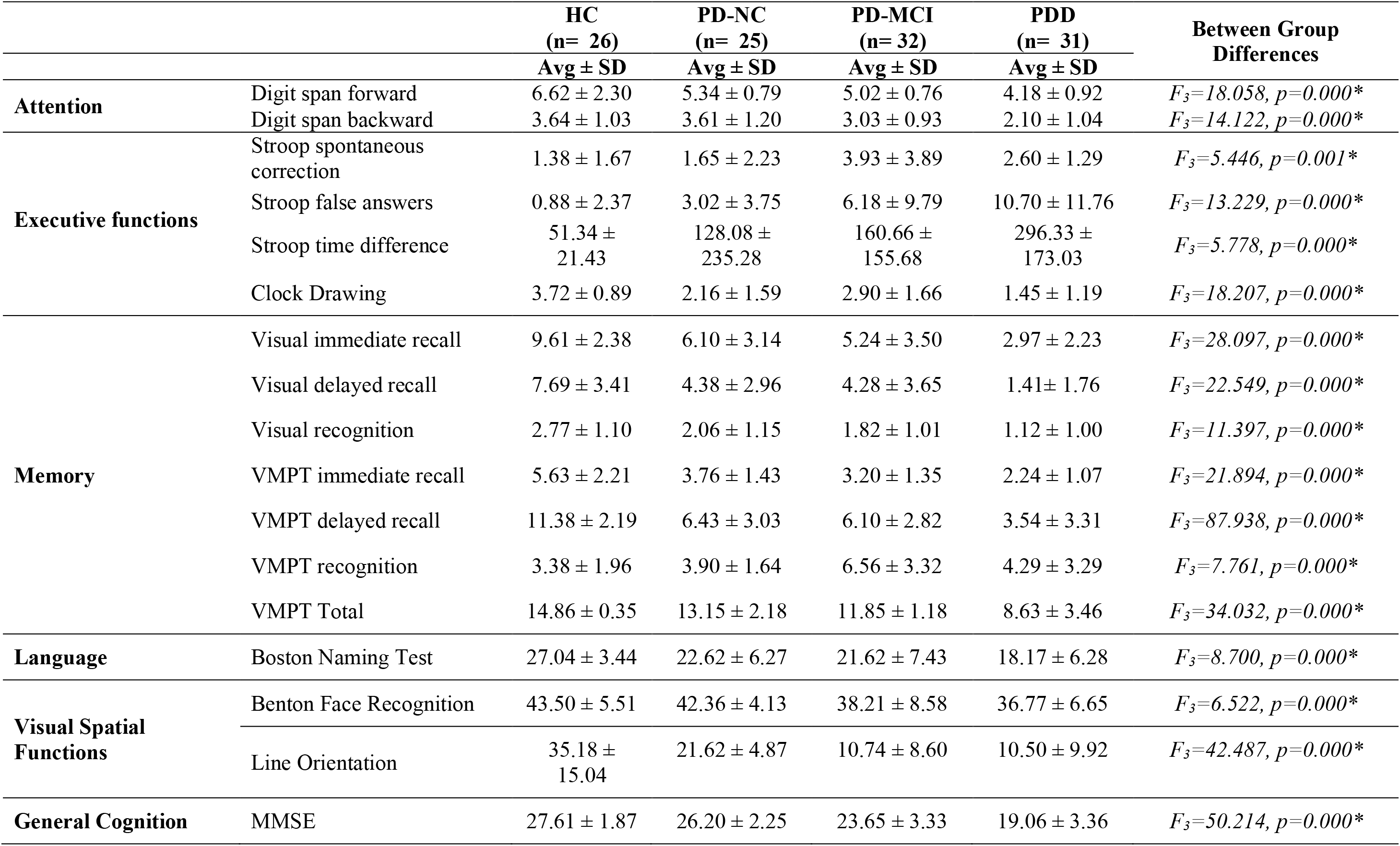
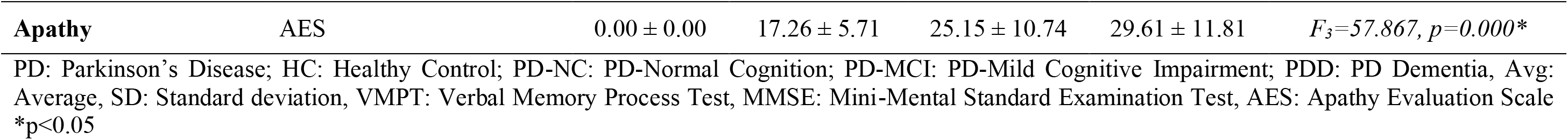
Clinical Between Groups Findings

**Figure 2.**
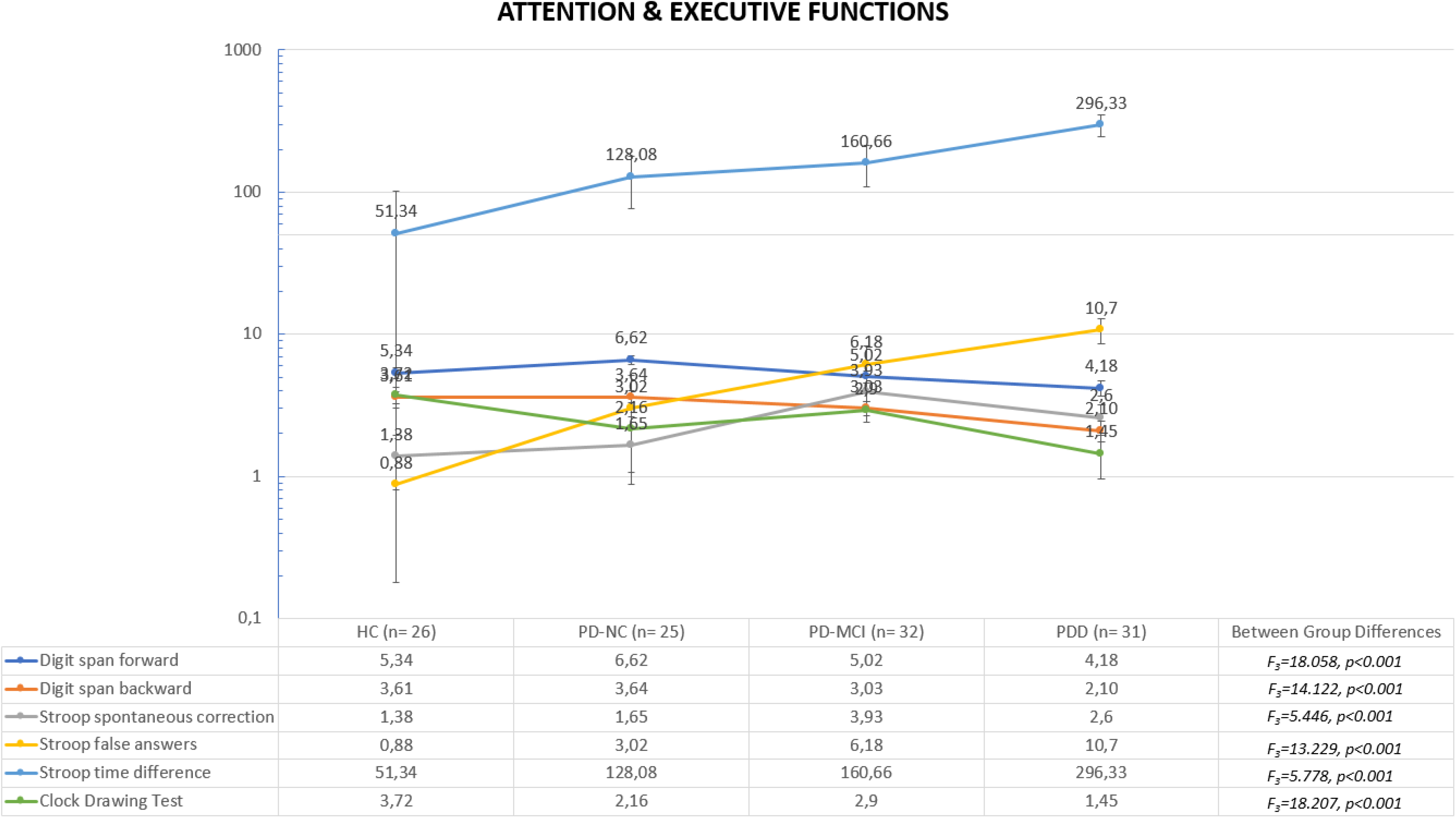
Clinical data findings of cognitive status – attention & executive functions

**Figure 2.**
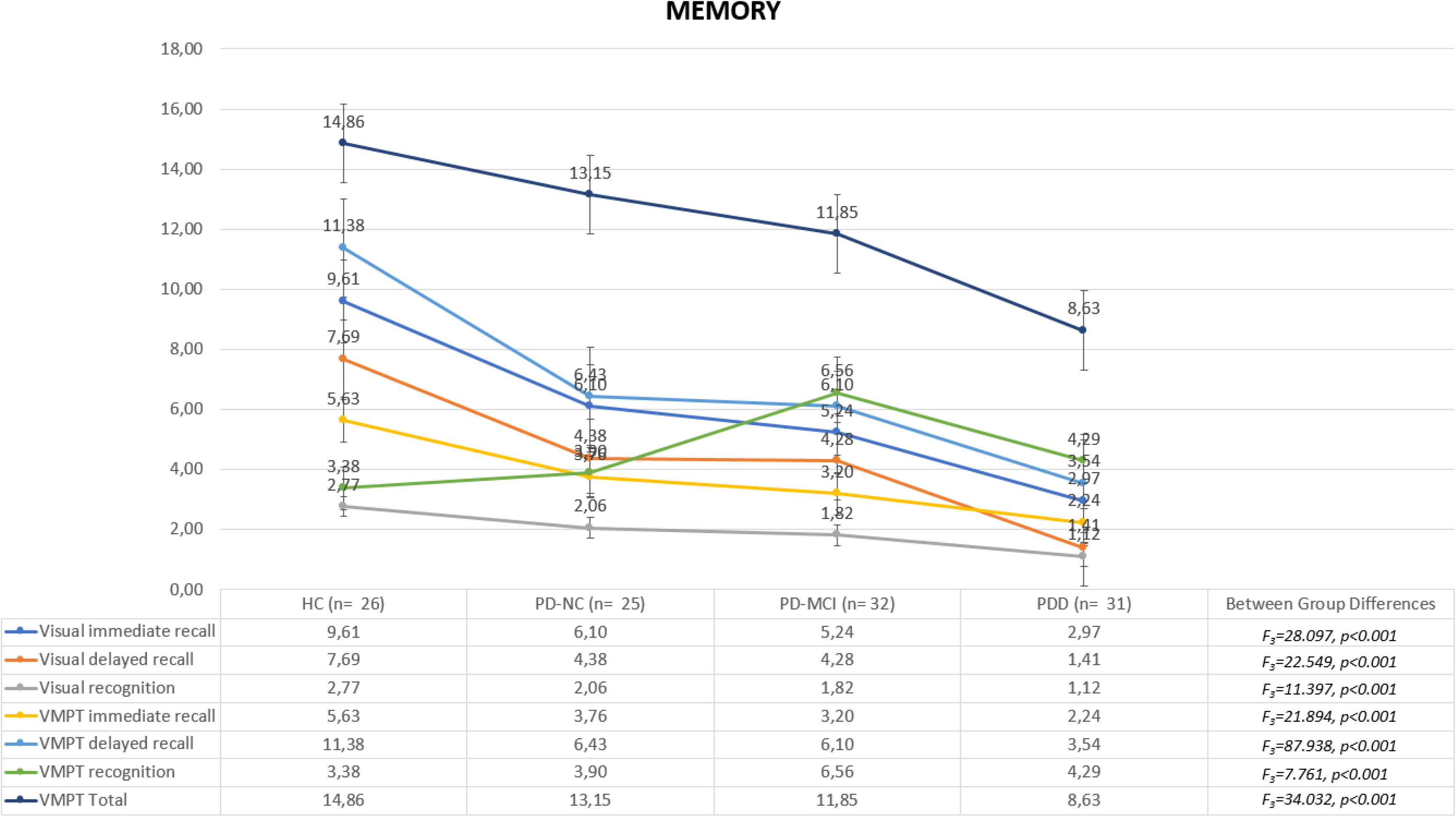
Clinical data findings of cognitive status -- memory

**Figure 2.**
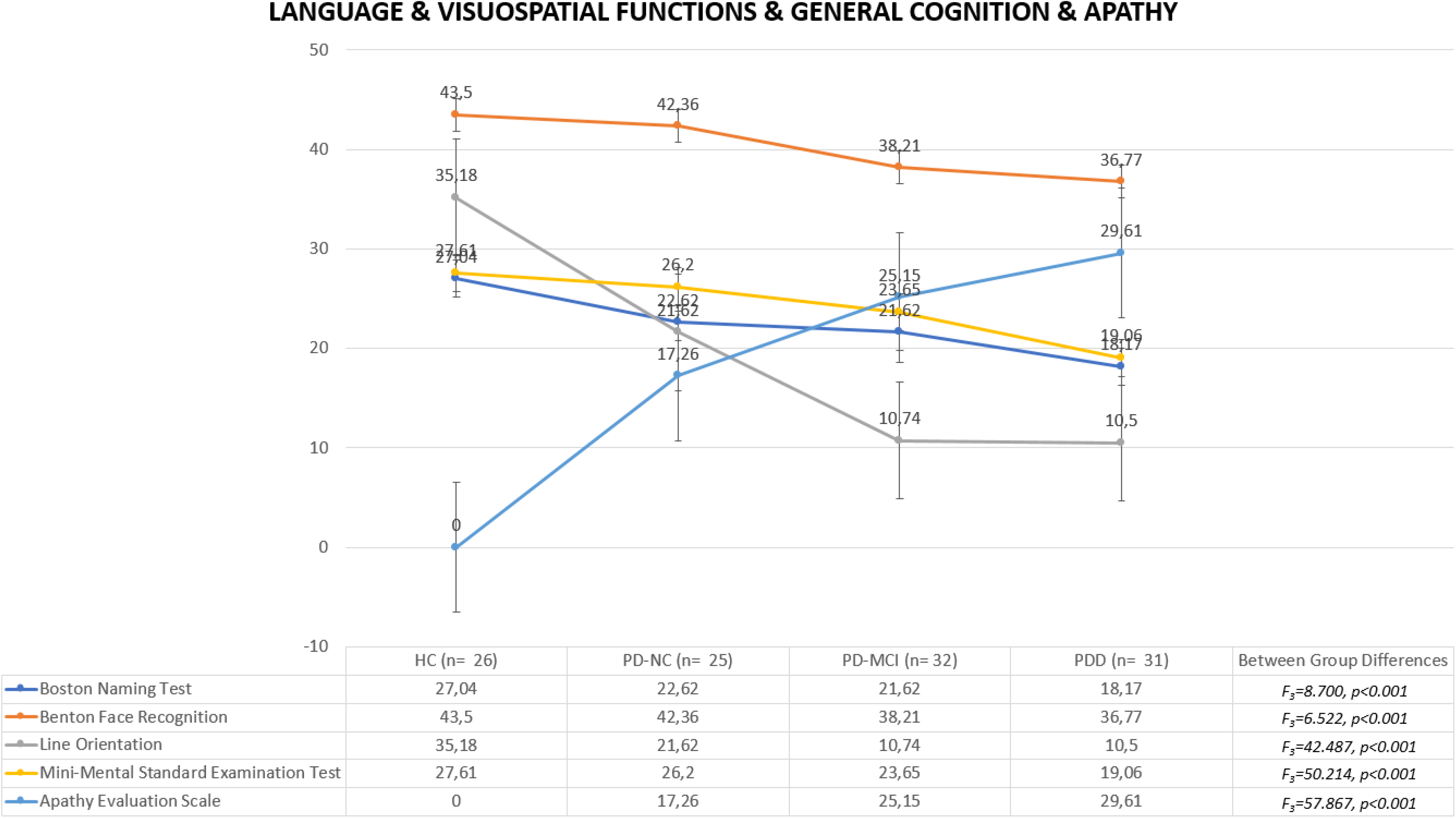
Clinical data findings of cognitive status – language & visuospatial functions & general cognition & apathy

## Functional Neuroimaging Results

### Group Comparison with Dual Regression

We conducted the final analysis with the data from 114 participants (88 PD patients and 26 HC), divided into four groups. We labeled 12 out of the 20 components as components of interest regarding their representative networks (Smith et al., 2009). Significant between-group differences in the network components and representative maps are given in Fig. 3 (p<0.05; corrected for multiple comparisons with threshold-free cluster enhancement (TFCE). We showed the comparison between groups as increased and decreased FC in each of the components, considering the transition of the disease continuum from HC to PD-NC, PD-MCI and PDD. Additionally, the number of voxels in the significant contrasts with minimum p values, t values, peak MNI152 coordinates, and their labels on the Harvard-Oxford Atlas are shown in Table 3. In general, our Dual Regression results showed that patients with PD-NC and PD-MCI had reduced FC in the left FPN compared to HCs. Additionally, decreased FC of DMN, left FPN, and SN and increased FC of SMN and VN were observed in PDD patients compared to HC. Briefly, compared to HCs, FC in the LFPN decreases in all three CI subtypes. Contrastingly, the PDD patients showed an additional decrease in the FC in the DMN and SN, while these patients had increased FC in the VN and SMN compared to HC. This increase in FC in SMN and VN was also seen in the comparison of PDD versus PD-NC. In comparison of PDD versus PD-MCI, we found decreased FC of the DMN, Temporoparietal Network, and VN and increased FC of DMN and VN. Therefore, different nodes of DMN and VN in PDD showed both increased and decreased FC when compared to PD-MCI.

**Figure 3.**
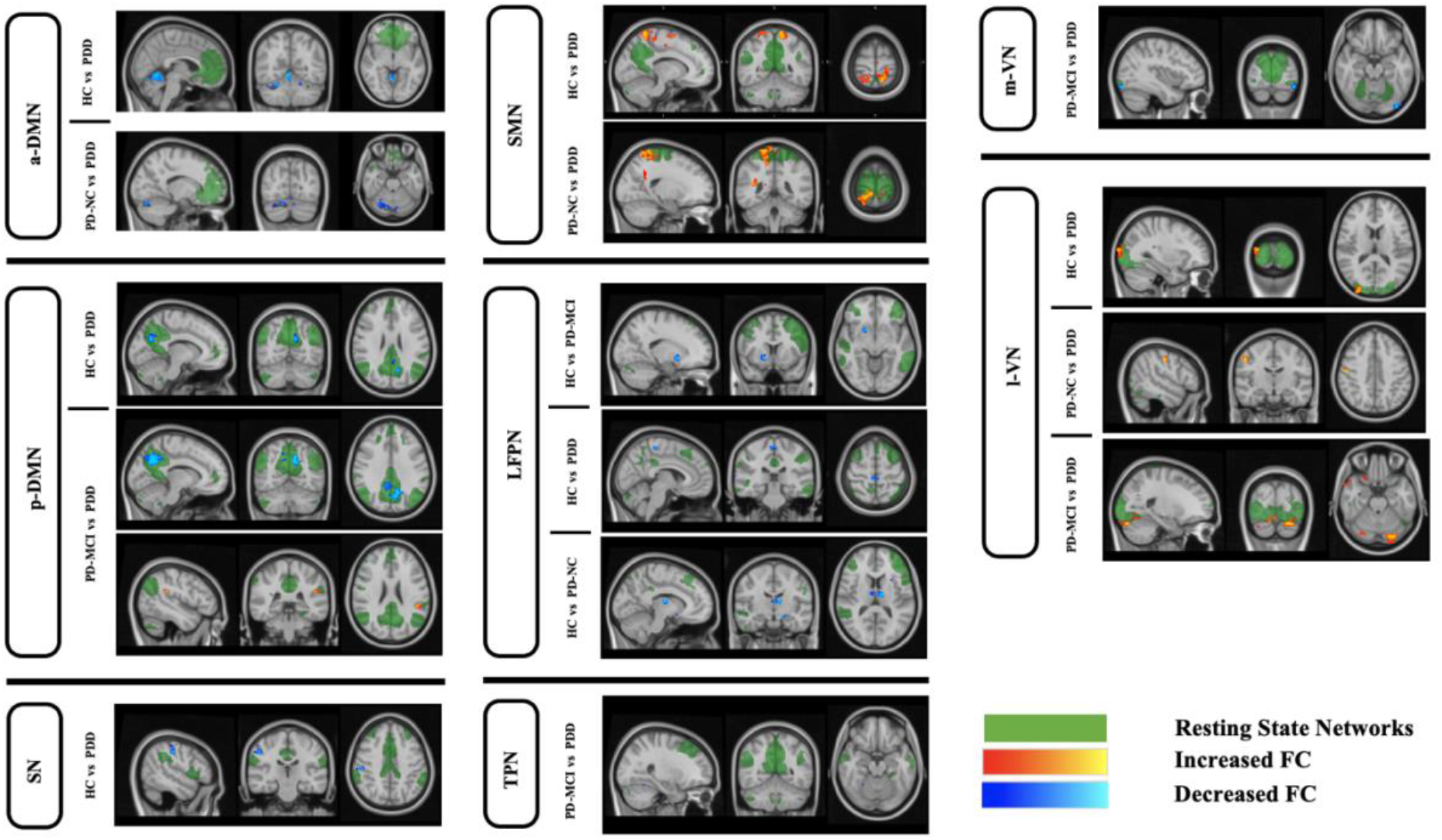
- Significant results of group-level ICA between groups p<0.05; corrected for multiple comparisons with Threshold-Free Cluster Enhancement (TFCE). PD: Parkinson’s Disease; HC: Healthy Control; PD-NC: PD-Normal Cognition; PD-MCI: PD-Mild Cognitive Impairment; PDD: PD-Dementia; a-DMN: Anterior Default Mode Network; p-DMN: Posterior Default Mode Network LFPN: Left Frontoparietal Network; SMN: Sensory-Motor Network; SN: Salience Network; TPN: Temporoparietal Network; mVN: Medial Visual Network; lVN: Lateral Visual Network. More information on the anatomical locations is given in Table 3.

**Table 3.**
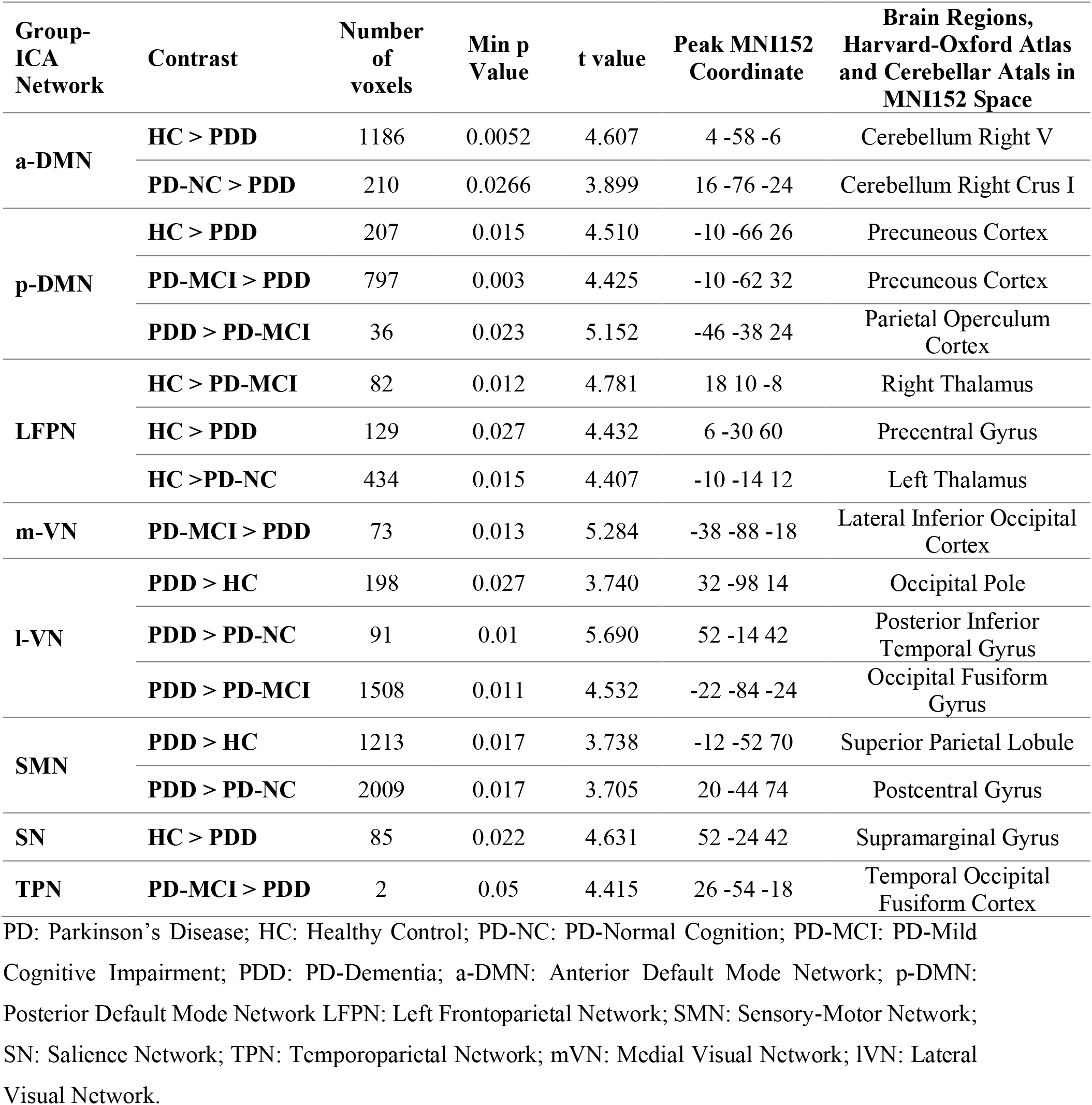
Between Groups Comparisons, Results of group-level ICA, Corrected for Multiple Comparisons with Threshold-Free Cluster Enhancement (TFCE, p < 0.05)

### Network Connectivity with FSLNets

Four hierarchical clustering dendrograms were calculated and presented as HC, PD-NC, PD-MCI, PDD (Supplementary File 1). These dendrograms can show the progression of sparseness from HC toward PD and among CI subtypes, from PD-NC to PD-MCI and PDD. Group comparisons regarding network connectivity based on Schaefer parcellation (accepted as significant at a p<0.05 FWE corrected for multiple comparisons, TFCE), revealed significant differences in various contrasts between 10 pairs of nodes. The edges between these significant nodes and related boxplots showing edge strength differences between groups are shown in Table 4 and Fig. 4. Nodes that were significant more than once (at least 2) were used to visualize the difference between the groups regarding the inter-network connectivity. Nodes 35 (Left Precuneus), 39 (Left Posterior Cingulate Cortex), 91 (Right Posterior Cingulate Cortex), and 27 (Right Medial Posterior Prefrontal Cortex) were significant more than once, and their connection in the connectogram is used for visualization purposes in the inter-network comparisons (Fig. 5). Finally, in Fig. 6, the continuum of the disease was shown to better represent the differences between groups and show the transition in the CI subtypes of PD. Overall, our network analysis with FSLNETS revealed that, compared to HC, FC between nodes of the DMN-VN in PD-MCI increased, while FC between nodes of the DMN in PDD decreased. In addition, increased FC of SN-DMN was observed in PD-MCI compared to PD-NC. Importantly, decreased FC of FPN and DMN (both correlation and anti-correlation) was observed in their own nodes and also in communication with each other in PDD compared to PD-MCI. These results admitted the positive and negative synchronizations in PDD compared to the PD-MCI. Finally, these altered synchronizations were accompanied by a decreased FC of VN-DAN in PDD compared to PD-MCI.

**Table 4.**
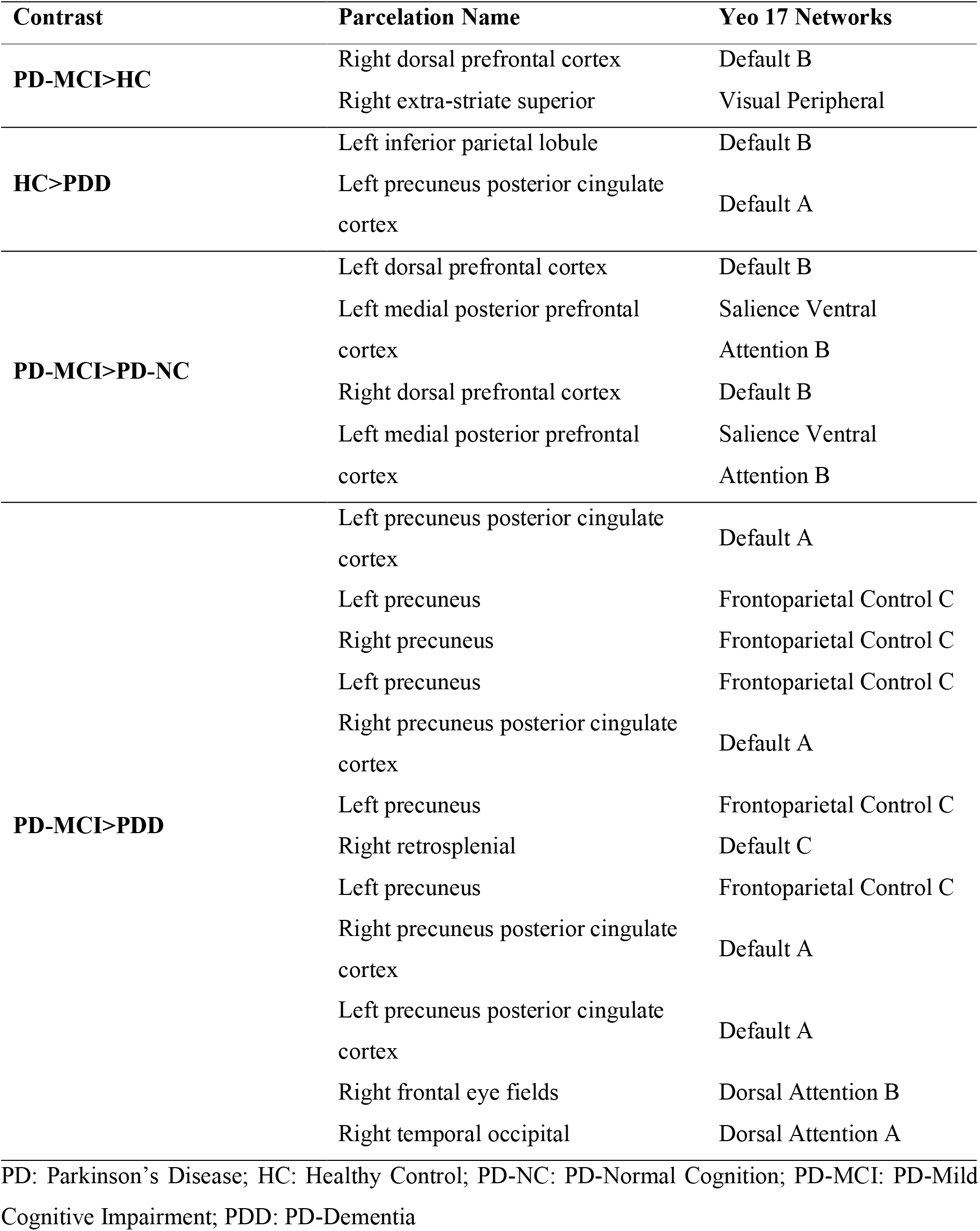
-Inter-network differences between groups, Schaefer100 Parcellation. Results of non-parametric tests using 5000 random permutations, corrected for multiple comparisons with Threshold-Free Cluster Enhancement (TFCE, P-value < 0.05).

**Figure 4.**
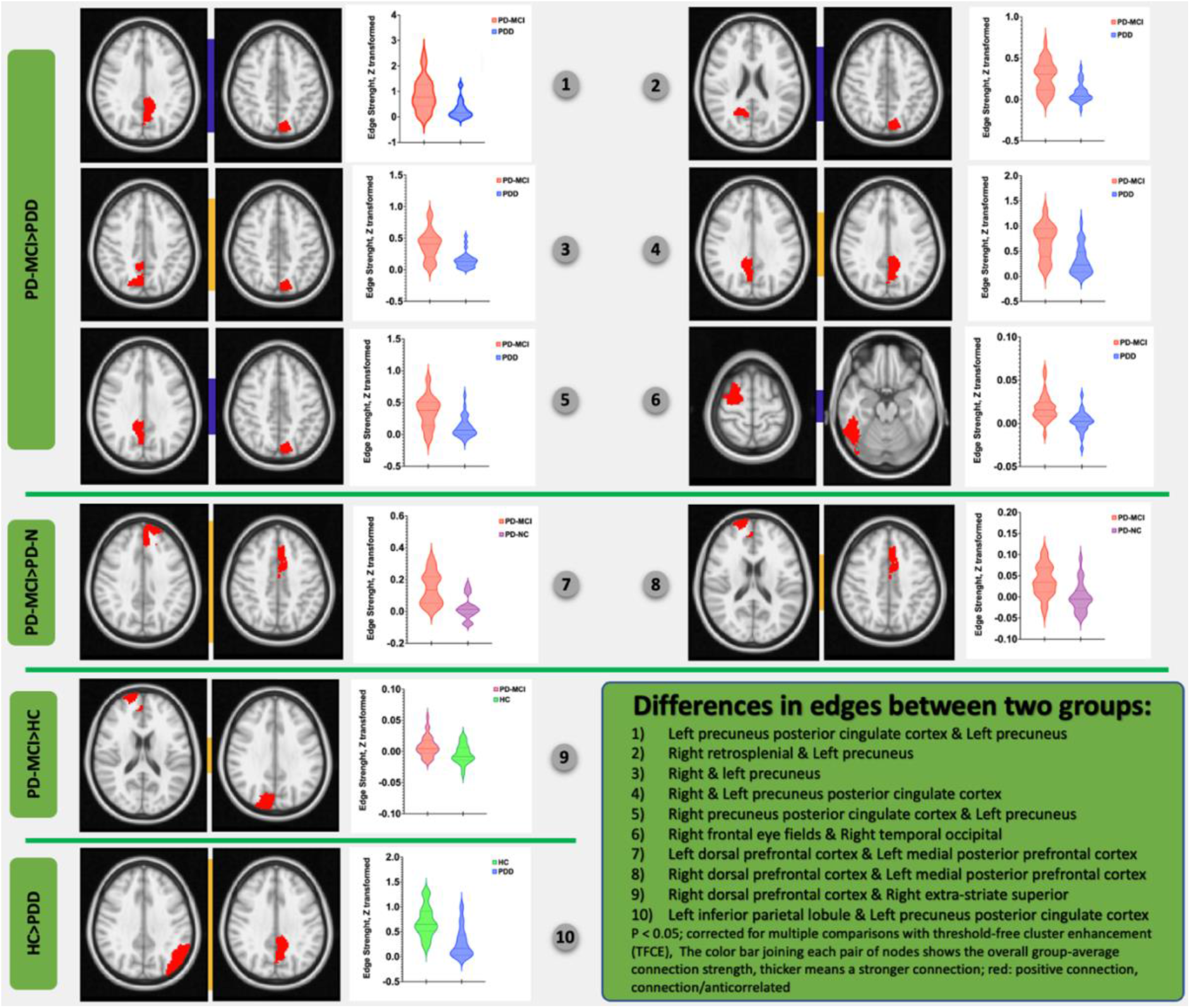
- Significant differences in Edge Strength (Z transformed) across every two groups based on Shaefer 100 parcellation, showing significant differences in the connectivity between two nodes and related boxplots (p<0.05, FWE-corrected). More information on the anatomical locations is given in Table 4. PD: Parkinson’s Disease; HC: Healthy Control; PD-NC: PD-Normal Cognition; PD-MCI: PD-Mild Cognitive Impairment; PDD: PD-Dementia

**Figure 5.**
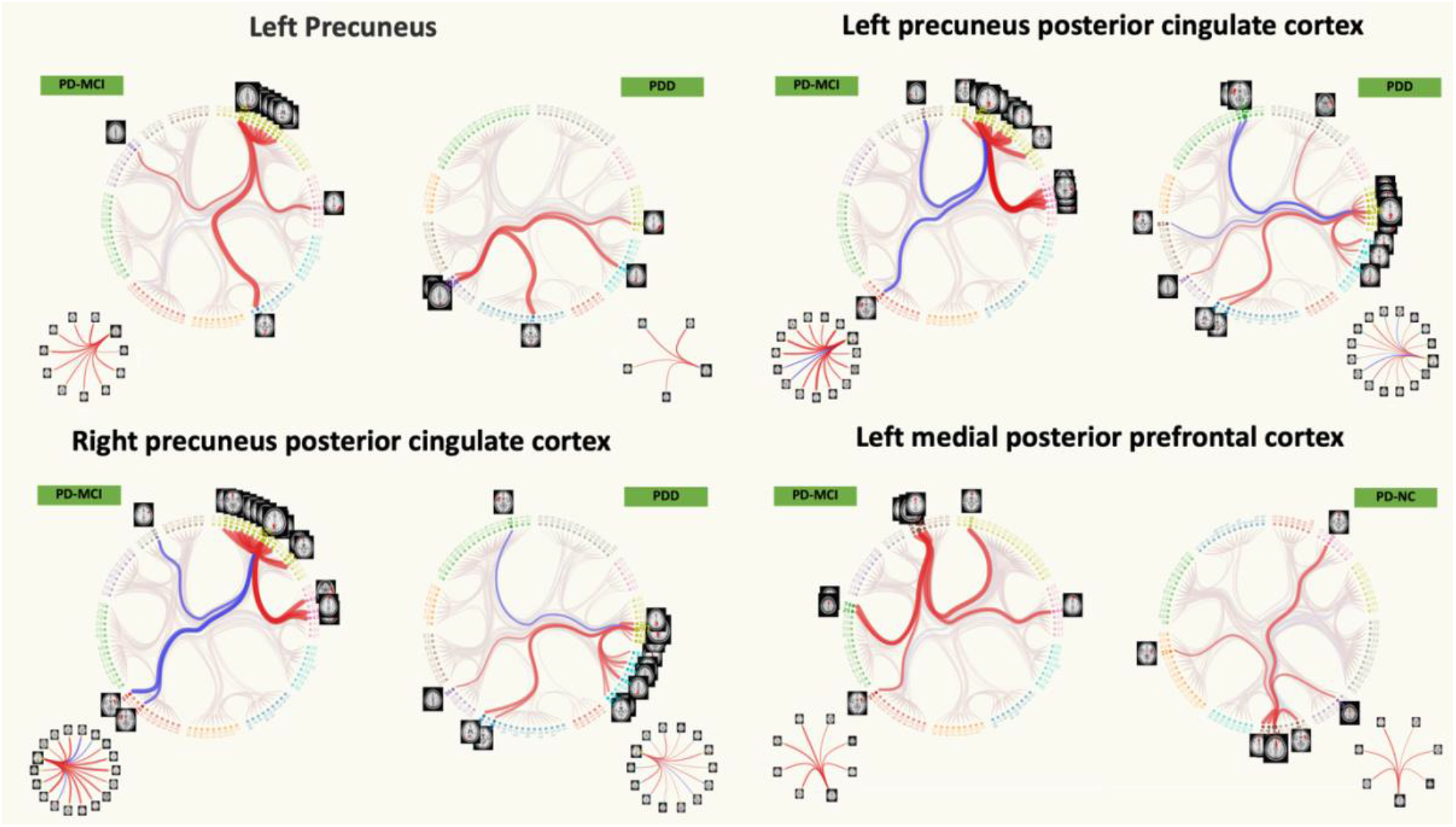
- Differences between connectograms in the inter-network comparisons. Red connections (edges) show positive correlations, and blue connections show anti-correlations.

**Figure 6.**
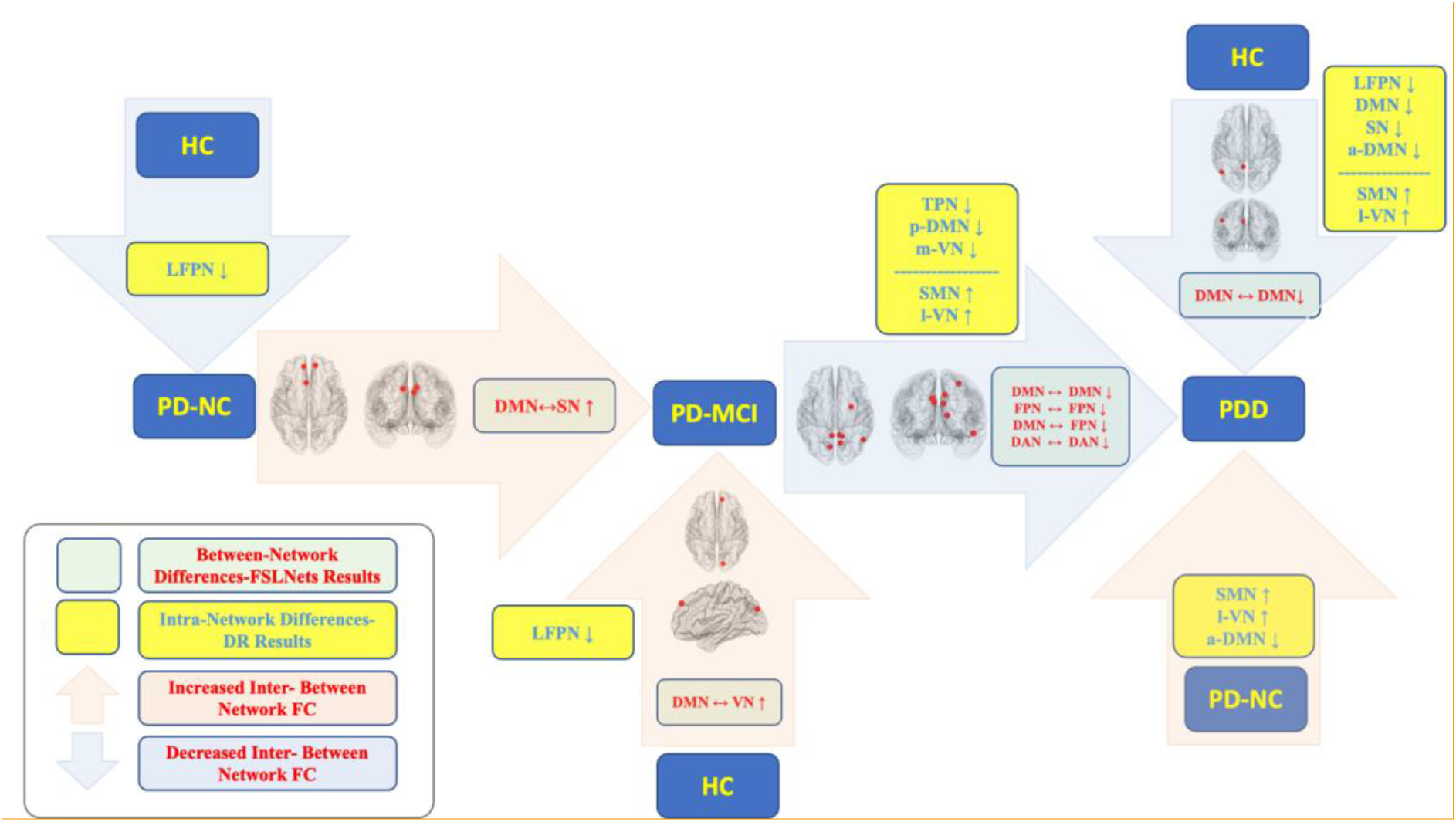
- The continuum map of FC changes in CI subtypes of the PD

## Discussion

Using clinical behavioral data and rs-fMRI data of patients with PD, we showed the differences between three subtypes of PD related to the cognitive phenotype, including PD-NC, PD-MCI, and PDD, and compared them with HC. Hypothetically, PD progresses from stages without CI to MCI and to dementia. PD-MCI is a transitional stage that comes after PD-NC and before PDD and can be a sign of dementia in people with PD (Yu & Wu, 2022). More challenges with daily living arise for PD patients as the disease progresses from PD-NC to PD-MCI or PDD (Leroi et al., 2012). Discovering the biomarkers linked to various cognitive PD phenotypes and, later, classifying PD patients into the appropriate cognitive subtype of PD will be crucial for planning the necessary pharmacological, rehabilitative, or neuromodulatory treatment plans. Particularly, neuroimaging results would play a pivotal role in separating out the various CI subtypes of PD (Devignes et al., 2022; Martín-Bastida et al., 2021). In a comprehensive review by Wolters et al., from 17 studies included, comparing the rs-fMRI results only two studies have reported differences between four groups (Wolters et al., 2019). In one of the studies, only 6 participants with PDD were included (Gorges, Müller, Lulé, LANDSCAPE Consortium, et al., 2015) who were evaluated together with other patients with CI in the same group; therefore, the results could not be distinguished between PD-MCI and PDD. In the other study with 27 PD patients, only 9 PDD patients are included in the study, and the whole hypothesis has been made on the PCC (Zhan et al., 2018). The conclusion of this review paper has been reported according to the evaluation of both the PD-MCI and PDD populations as one population of patients with CI. Furthermore, in an important study by Fiorenzato et al., 3 subtypes of PD have been investigated to show the differences in brain network connectivity only by utilizing the temporal dynamic changes (Fiorenzato et al., 2019). Here, our results demonstrate clinical and neuroimaging differences between CI subtypes of PD. We discuss the differences between each of the CI subtypes with HC and further review the differences between the CI subtypes of PD with each other.

### CI Subtypes of PD versus HC

Although defined as cognitively normal, a subtle cognitive impairment is also reported in the PD-NC population without the diagnosis of PD-MCI (Chua et al., 2021). Compared to HC, PD-NC has both increased and decreased FC, including decreased FC of FPN with DMN, VN, and DAN (Klobušiaková et al., 2019) and increased FC of VN-LFPN with a reduction in visuospatial processing by demonstrating a crucial decline in working memory ((Wei et al., 2022). In an important study by Peraza et al., both decreased and increased FC in PD-NC and increased FC in PD-MCI compared to HC, as well as increased between-network FC of basal and motor networks, are reported (Peraza et al., 2017). The increased FC/hyperconnectivity in the DMN, left and right FPN, SN, motor, basal ganglia-thalamic, and brainstem in patients without CI is interpreted as a compensation for the neuronal loss due to the pathophysiology of the PD. On the other hand, this hyperconnectivity has changed to hypoconnectivity in PD patients with PD-MCI mostly seen in the DMN, motor, and DAN. Therefore, while there can be both a preserved and increased FC of DMN in the early stages of the disease preceding PD-MCI, the introduction of CI may lead to the loosening of connections specifically at nodes of DMN (Gorges, Müller, Lulé, LANDSCAPE Consortium, et al., 2015). The above-described changes in FC can show heterogeneity in study results. As the disease progresses to the severe stages from PD-MCI to PDD, the FC and clinical outcomes continue to deteriorate. Over the course of a 3-year follow-up, PD patients show a progressive loss of FC in several brain regions, as confirmed by longitudinal studies (Dubbelink et al., 2014).

FPN and DMN are two important networks associated with CI in PD. The integration of cognitive and emotional processing, as well as mind-wandering, are considered to be regulated by the DMN. On the other hand, FPN is associated with higher cognitive skills such as actively preserving and processing information in working memory, problem solving and executive control, and making decisions in the context of goal-directed behavior (Grady et al., 2016). The impaired cognitive functions of PD, such as attention, executive functions, memory, and visuospatial abilities, can be associated with altered FC in related networks such as DMN and FPN due to the progressive neurodegenerative effect of the disease (Baggio et al., 2015; Dubbelink et al., 2014; Gorges, Müller, Lulé, LANDSCAPE Consortium, et al., 2015; Mak et al., 2015). Compared to HC, decreased FC of DMN was reported in PD-NC and PD-MCI, and decreased FC of FPN with bilateral PFC was reported in PD-MCI (Amboni et al., 2015). In advanced levels of the disease, subjects with PD-MCI and PDD demonstrate impaired FC in the FPN and its associated networks, even when controlled for dopaminergic medications (Borroni et al., 2015; Rektorova et al., 2012). Moreover, PDD patients have shown decreased FC of DMN and VN compared to HC in a previous study using 1.5T scanning (Rektorova et al., 2012).

We discovered decreased FC of the FPN in all three subtypes compared to HC. FPN is an important neurocognitive network that can play a role in all CI subtypes of PD. Altered FC of FPN in our study can be interpreted as deteriorated clinical outcomes in all three groups, such as executive functions, memory, and visuospatial functions. Substantially, our DR results in the PD-NC and PD-MCI groups were significant in the FC of the left FPN with thalamus. Reduced FC of the mediodorsal thalamus with the paracingulate gyrus has previously been linked to CI in PD (Owens-Walton et al., 2021). In our results, the decreased FC of FPN extended beyond the network (in the thalamus) and can be interpreted as decreased communication of FPN with the thalamus. While the FPN network is crucial for the cognitive abilities of PD patients, its association with the thalamus, one of the primary subcortical brain regions, can be significant. Additionally, our results showed decreased FC of the supramarginal gyrus in PDD versus HC. The results of an important neuroimaging meta-analysis by Tahmasian et al., have shown the importance of the supramarginal gyrus in PD patients compared to HC, which is affected by the role of dopamine replacement therapy, and highlight the importance of this region in the neuropathology of PD. Our results emphasize the importance of this region also in the advanced stages of the CI levels (Tahmasian et al., 2017). Our results in the PDD group compared to HC, showed reduced FC of FPN, DMN, and SN and increased FC of SMN and VN. This was admitted by our network analysis results as decreased interconnection in the posterior nodes of DMN was found in PDD compared to HC, while communication was increased in the FC of VN and DMN in PD-MCI compared to HC. This can acknowledge the fact that the gradual decreased FC of neurocognitive networks in the progression of CI in PD results in worse cognitive functions, but in the advanced stages of CI, the load on motor and visual networks may increase to compensate for this cognitive decline. Additionally, PD-MCI patients may try to increase the coupling between posterior-frontal regions of the brain (DLPFC and superior extrastriate cortex) that may be an indicator of a strategy in PD-MCI patients that try to rely on their imagery visualization to preserve their memory. Of note, differences between HC vs PD-MCI and PDD in our study may contribute to the previous knowledge in the literature that PD-MCI is more associated with the fronto-striatal dopaminergic dysfunction, which embraces clinical problems in executive functions and working memory, while PDD is associated with posterior cholinergic dysfunction that is involved in visuospatial dysfunctions and can be a predictor of worse cognitive progression (Kehagia et al., 2013).

### PD-NC vs PD-MCI & PDD

The most targeted intervention and care should be used to prevent PD patients from progressing to advanced CI. Therefore, comparing CI subtypes of PD with each other and investigating their progression from PD-NC is important. Many studies have shown decreased inter and intra-network FC in various networks in the progression from PD-NC to PD-MCI, predicting the CI (Amboni et al., 2015; Gorges, Müller, Lulé, Pinkhardt, et al., 2015; Lopes et al., 2017; Peraza et al., 2017), as well as a reduction in normal anticorrelation in DMN-DAN (Baggio et al., 2015). Like in comparisons to HC explained above, increased FC in the preceding stage before progression of the CI has given its place to hypoconnectivity in patients with CI, specifically MCI (Gratton et al., 2018). A recent longitudinal study showed that decreased FC between two nodes of the DMN (mPFC and PCC) can be a predictive factor in the conversion from PD-NC to PD-MCI (Zarifkar et al., 2021). Additionally, with progression of the PD, FC within SMN and the interconnection between DAN and FPN decreases and progressive decline in the DAN-PFN FC is associated with the CI in PD, that can further be used as a marker in assessing the progression into PDD (Campbell et al., 2020).

However, some studies have also shown increased FC in the progression from PD-NC. Increased FC in cerebellar and insular networks (Peraza et al., 2017) and hyperconnectivity of PCC with several frontal and posterior regions are reported in PD-MCI compared to PD-NC (Zhan et al., 2018). Results of a graph theory study showed decreased long range connectivity but increased local interconnectedness in PD-MCI compared to PD-NC which was linked to visuospatial and memory functions (Baggio et al., 2014). In two studies by Aracil-Bolaños et al., stronger functional coupling between nodes of the normally anticorrelated DMN and Central Executive Network, as well as increased SN-DMN FC, are reported in PD-MCI compared to PD-NC (Aracil-Bolaños et al., 2019, 2022). With the deterioration of cognition in the progression of PD, FC changes as well. Reduced FC of DMN in the right inferior frontal gyrus in PDD compared to PD-NC has been shown in a previous study using 1.5T scanning (Rektorova et al., 2012). The anterior cingulate cortex, caudate nucleus, medial and dorsolateral prefrontal cortices, and left precentral gyrus are recruited more strongly in PDD and PD-MCI patients than in PD-NC patients during working memory or executive function tasks (Nagano-Saito et al., 2014).

Our clinical and network analysis can admit the worse visuospatial functions in PDD and PD-MCI compared to PD-NC. Moreover, our results supported the increased FC between DMN and SN, as close nodes in the frontal regions of the brain showed increased FC in PD-MCI compared to PD-NC. While less is reported about the comparisons between PD-NC and PDD in the literature, we observed increased FC in SMN and VN, probably as a compensation mechanism, with regard to decreases in the FC of the neurocognitive networks in PDD compared to PD-NC. Therefore, from PD-NC to PD-MCI and PDD, communication between close regions of the brain increases with the communication between DMN and SN in PD-MCI, and later in the PDD level that the decline in neurocognitive networks is settled, the terminal load is seen on SMN and VN to compensate the functional loss associated with PDD. Further studies may be needed to support this hypothesis that from PD-NC to MCI or PDD, the increased FC between neurocognitive networks such as DMN-SN can be a predictor of PD-MCI, but increased FC in the SMN and VN can be a predictor of the PDD commence, therefore needed for emergent action. All this information should be supported by the clinical neuropsychological data.

### PD-MCI vs PDD

Differentiating between PD-MCI and PDD can be the most critical step in differentiating the CI subtypes of PD, as dementia can have debilitating effects on patients with an increased hospitalization rate and related life difficulties. Therefore, when possible, early detection and the application of effective interventions can be highly beneficial for patients with PD (Biundo et al., 2016). Longitudinal studies have shown a progressive age-independent decrease in FC in posterior brain regions-parietotemporal that may predict the CI towards PDD in patients with PD-MCI (Dubbelink et al., 2014). Previous studies have also shown that the FC between the frontal cortex and the corticostriatal cortex is disrupted in PDD, which leads to cognitive decline (Rektorova et al., 2012; Seibert et al., 2012). From PD-MCI to PDD, a decrease in FC of PCC is found with subcortical nuclei including the left caudate and right thalamus, as well as cortical regions including the precuneus, middle frontal, and right angular gyri in PDD compared to PD-MCI. Additionally, a compensatory loop connection is introduced between the cerebellum and frontoparietal networks in PD with CI (Zhan et al., 2018). Increased FC in the left middle and superior frontal gyri remains the same when comparing PDD to PD-NC. However, the FC between the PCC and anterior cingulate and paracingulate gyri changes to hypoconnectivity in PDD compared to PD-NC (Zhan et al., 2018). It is important to note that the described work by Zahn et al., was performed with only 9 participants in each group. It can be inferred from this study that PCC, which is one of the main hubs in DMN, plays a key role in the progressive CI from PD-NC to PDD, as its FC increases from NC to MCI and then decreases from MCI to PDD. Similarly, two nodes of the DMN, PCC, and middle prefrontal cortex, showed increased FC in the absence of CI (PD-NC) and in the PDD (Chen et al., 2015). Regarding clinical symptoms, although memory performance is a significant determinant of the development of PDD, other cognitive processes like attention, executive function, visuospatial function, and language also play an important role in PDD development (Galtier et al., 2016). Biundo et al. concluded that language and executive functions, together with visuo-spatial and visuo-perceptual abilities, demonstrated the best sensitivity in detecting PDD (Rektorova et al., 2014). Our clinical data showed significant impairments in terms of attention, executive function, memory, and general cognition in PDD versus PD-MCI. Patients with PDD had more significant cognitive impairments than PD-MCI patients, particularly in the frontal/executive and memory domains. These findings may have consequences for cognitive prognosis. Our results showed that moving from PD-MCI to PDD can lead to decrease in DMN-FPN and DAN-VN functional connections. Importantly, our results were most dominant in the DMN-FPN. Therefore, it can be inferred that not only is the connectivity within the DMN and FPN important in differentiating the dementia levels of PD patients, but how these two networks communicate with each other is also important in describing the progression from PD-MCI to PDD. We also found both increased and decreased FC in DMN and VN of PDD compared to PD-MCI. Therefore, we can propose that DMN and VN can show different changes in FC with other brain regions when transitioning from PD-MCI to PDD. This can further be supported by the decreased FC inside DMN, FPN, and DAN (the inter-network communications inside these networks decrease) and also between each other (both DMN and FPN communicate less with each other). Although decreased and increased FC of DMN and VN can be seen in the transition from PD-MCI to PDD, how DMN communicates in its own nodes and especially with FPN, and whether there’s a hypofunction in the nodes of DAN, can be a classifier in further studies to capture PDD after PD-MCI.

### Limitations

Our study has some limitations. First, our results do not contain any longitudinal information. As Parkinson’s disease is a progressive disease, evaluating the longitudinal data from the same population would give more overwhelming information regarding the progression of the disease and continuum of cognitive impairment. Second, there were statistically significant differences in demographic parameters, and although we tried to match the patient groups with HC and included age as a covariate of no interest in our data analysis, patients were significantly older than the HCs.

### Conclusion

Our results showed the importance of DMN, FPN, DAN, and VN and their inter-intra network FC in distinguishing the difference between PD-MCI and PDD. Additionally, our results showed the importance of SMN, VN, DMN, and SN in the progression from PD-NC to PDD. In comparison to HC, we found DMN, FPN, VN, and SN as important networks for further differential diagnosis of CI subtypes of PD. With emergence and progression of the diseases, patients with PD rely on different strategies in several networks to compensate for the loss in FC of neurocognitive networks, specially DMN and FPN. We propose that, if approved with clinical data, from PD-NC to MCI or PDD, the increased Functional coupling between neurocognitive networks such as DMN-SN can be a predictor of PD-MCI, but increased FC in the VN and SMN can be a predictor of the PDD. We further propose that although decreased and increased in DMN and VN can be seen in transition from PD-MCI to PDD, but how DMN behaves in the inter- and intra-network level specially with FPN, and a hypofunction in nodes of DAN can be a predictor factor in further studies to capture PDD after PD-MCI. All these results were in line with clinical changes in cognitive functions. Utilizing clinical and neuroimaging data with machine learning and artificial intelligence may be capable of capturing early stages of PD in clinical settings to better plan the interventions in PD and consequently prevent progression of PD to more advanced levels of the disease.

## Data Availability

The data that support the findings of this study are available from the corresponding author upon reasonable request.

## Data Availability

All data produced in the present study are available upon reasonable request to the authors

## Acknowledgements

The authors acknowledge all the participants who accepted to take part in this study. We also thank Dr. Nesrin H. Yılmaz, Dr. Zeynep T. Yıldız and Dr. Tuğçe Kahraman.

---

**Supplementary File 1.**
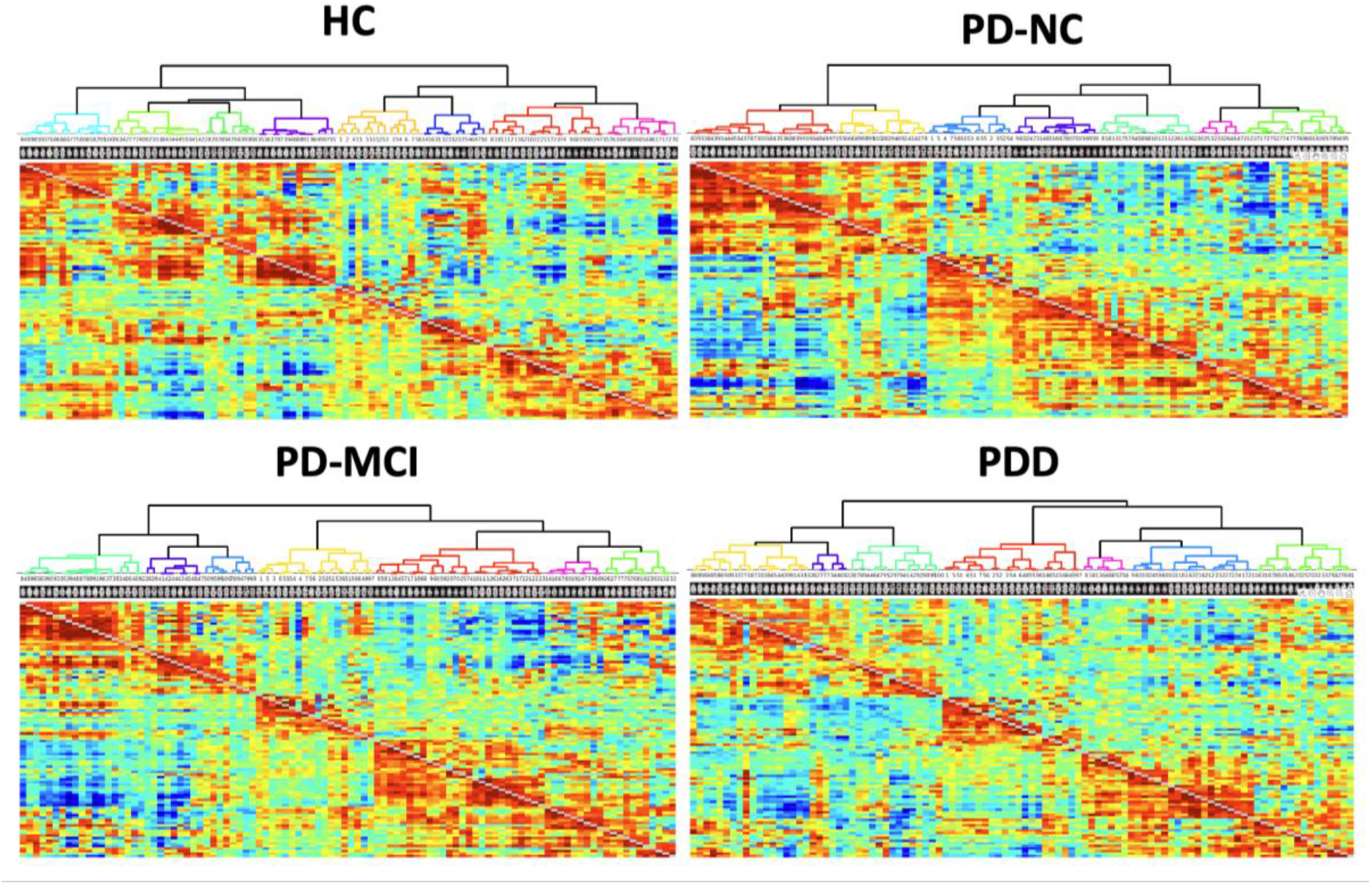
- Hierarchical Clustering Dendrogram in 4 groups (see methods, Inter-Network Connectivity Analysis), coloring in the dendrogram indicates main clusters with high temporal correlations thresholded at 0.75, as a default value in nets_hierarchy code of FLSNets. The upper and lower triangular matrices represent the partial and full correlations in all the dendrograms.

